# Preconception environments shape pregnancy-related structural brain remodeling and intergenerational mental health outcomes

**DOI:** 10.64898/2026.04.28.26351998

**Authors:** Xing Qian, Shi Yu Chan, Wen Liang Loh, Anne Rifkin-Graboi, Johan G. Eriksson, Marielle Valerie Fortier, Yap Seng Chong, Ai Peng Tan, Juan Helen Zhou

## Abstract

Pregnancy is accompanied by structural brain remodeling in networks supporting socio-emotional processing and cognitive control. However, it remains unclear whether and how modifiable preconception environments shape individual variability in pregnancy-related brain remodeling, and whether such environment-shaped brain features relate to subsequent maternal and offspring mental health. Here we leverage the Singaporean S-PRESTO preconception cohort, with brain structural MRI acquired before conception (PCV; n = 194) and at 3 months postpartum (PNV; n = 61). Age-related gray matter volume (GMV) trajectories were modeled based on PCV data to derive longitudinal region-wise deviation scores, accounting for normative age trends. Pregnancy-related GMV reductions were found in default mode, frontoparietal control, salience, and limbic networks, as well as hippocampus and caudate. We then identified a multivariate pattern linking preconception lifestyle, sociodemographic, psychological, and social-support variables to GMV within these pregnancy-vulnerable brain regions using partial least squares approach. Specifically, higher preconception brain GMV was associated with more advantaged socioeconomic status, healthier diet, better sleep, non-smoking, stronger social support, and higher conscientiousness/lower neuroticism. This environment-associated brain pattern relates to better maternal executive function, lower depression, anxiety, stress and more favorable metabolic health. Importantly, the same brain resilience pattern also predicted lower postpartum anxiety and fewer offspring internalizing symptoms at age four. Together, these findings implicate modifiable preconception environments in shaping brain structural resilience to pregnancy-related remodelling and highlight preconception as a potential window to promote maternal brain health and positive intergenerational mental health outcomes.

## INTRODUCTION

Pregnancy induces one of the most profound biological transitions in a woman’s life, involving large-scale endocrine, immune, metabolic, and neural adaptations that support fetal development and prepare the women for caregiving [1]. This transition also requires substantial psychological adjustment and may represent a period of heightened vulnerability to mental health difficulties [2]. While up to 85% of women experience transient affective symptoms (“baby blues”) during the perinatal period, approximately 10-20% develop clinically significant and persistent conditions such as postpartum depression and anxiety disorders [3]. In addition, an estimated 80% of women experience subjective or objective cognitive difficulties across the transition to motherhood, including reductions in memory, concentration, and attentional control [4].

Emerging neuroimaging research demonstrates that pregnancy is accompanied by significant alterations in the maternal brain [4]. Converging evidence shows systematic reductions in gray matter volume (GMV), particularly in regions supporting social cognition and higher-order thinking [5–9]. These reductions are often interpreted as reflecting synaptic pruning and large-scale neuroplastic remodeling that may enhance neural efficiency for maternal caregiving [5, 10, 11]. However, findings from both pregnancy and non-pregnancy literature suggest that GMV reductions in specific regions may also confer emotional or cognitive vulnerability, indicating that these changes are not uniformly adaptive [5, 12–15]. Remarkably, pregnancy-related GMV alterations do not fully recover [7]; they persist for at least six years postpartum and may even remain detectable decades later, underscoring the enduring nature of this transformative biological event [5, 16, 17].

Despite these robust findings, women enter pregnancy with diverse backgrounds and experiences. Lifestyle and environmental exposures, including diet, sleep quality, substance use, physical activity, social support, and socioeconomic status, are known to influence brain structure, neuroplasticity, allostatic load, and stress reactivity [18–20]. Yet no study has comprehensively examined whether these modifiable preconception factors shape a woman’s neural resilience or vulnerability in brain regions that later undergo pregnancy-related structural changes.

Beyond maternal neural health, pregnancy-related brain changes may have downstream effects on maternal cognition, emotional well-being, and offspring socio-emotional development [11, 21, 22]. The brain circuitry underlying core maternal functions—such as emotion regulation, attention, reward processing, and caregiving—relies on limbic, prefrontal, and striatal pathways that are also central to psychiatric vulnerability, including anxiety, depression, and heightened stress sensitivity [23–26]. Maternal psychological functioning, in turn, powerfully shapes children’s early environments by influencing attachment quality, exposure to stress, and risk for emotional and behavioral difficulties [8, 27, 28]. However, no prior study has examined whether individual differences in grey matter volumes within pregnancy-vulnerable brain regions, particularly those associated with modifiable lifestyle and environmental factors, predict maternal cognitive and mental health trajectories across the perinatal period, nor whether such neural markers carry intergenerational significance for children’s socio-emotional development.

To address these gaps, we leveraged longitudinal neuroimaging and phenotypic data from the Singapore PREconception Study of long-Term maternal and child Outcomes (S-PRESTO) [29], a unique cohort in which women were scanned twice (before conception and postpartum), with their children followed into early childhood. Here, we asked whether and how modifiable preconception environments shape variability in pregnancy-related brain remodeling, and whether such environment-linked neural patterns are associated with subsequent maternal and offspring outcomes. We hypothesized that more favorable preconception environments would be associated with greater structural integrity in brain regions undergoing pregnancy-related change, and that this resilience pattern would predict better maternal and offspring cognition and mental health outcomes.

## RESULTS

This longitudinal study included women who underwent MRI scans at the preconception visit (PCV) and/or at the postnatal visit (PNV; 3 months postpartum, M03) (**Fig. 1**). After image quality control (see **Image Preprocessing**), a total of 194 women at PCV (mean age 31.09 ± 3.66 years; range 19.49 - 43.58; ethnicity: 133 Chinese, 33 Malay, 20 Indian, 8 Other) and 61 women at PNV (31.82 ± 3.92 years; range 19.81 - 42.21; 42 Chinese, 14 Malay, 3 Indian, 2 Other) with high-quality T1-weighted MRI scans were included, among whom 35 participated at both visits (30.33 ± 3.50 years; range 21.37 - 38.64; 25 Chinese, 8 Malay, 2 Other). To assess intergenerational relevance, we additionally examined children born to these participants. Child behavioral data at age four were available for 40 offspring of PCV mothers (4.05 ± 0.10 years; range 3.89–4.40) and 52 offspring of PNV (M03) mothers (4.05 ± 0.08 years; range 3.89–4.31).

**Figure 1.**
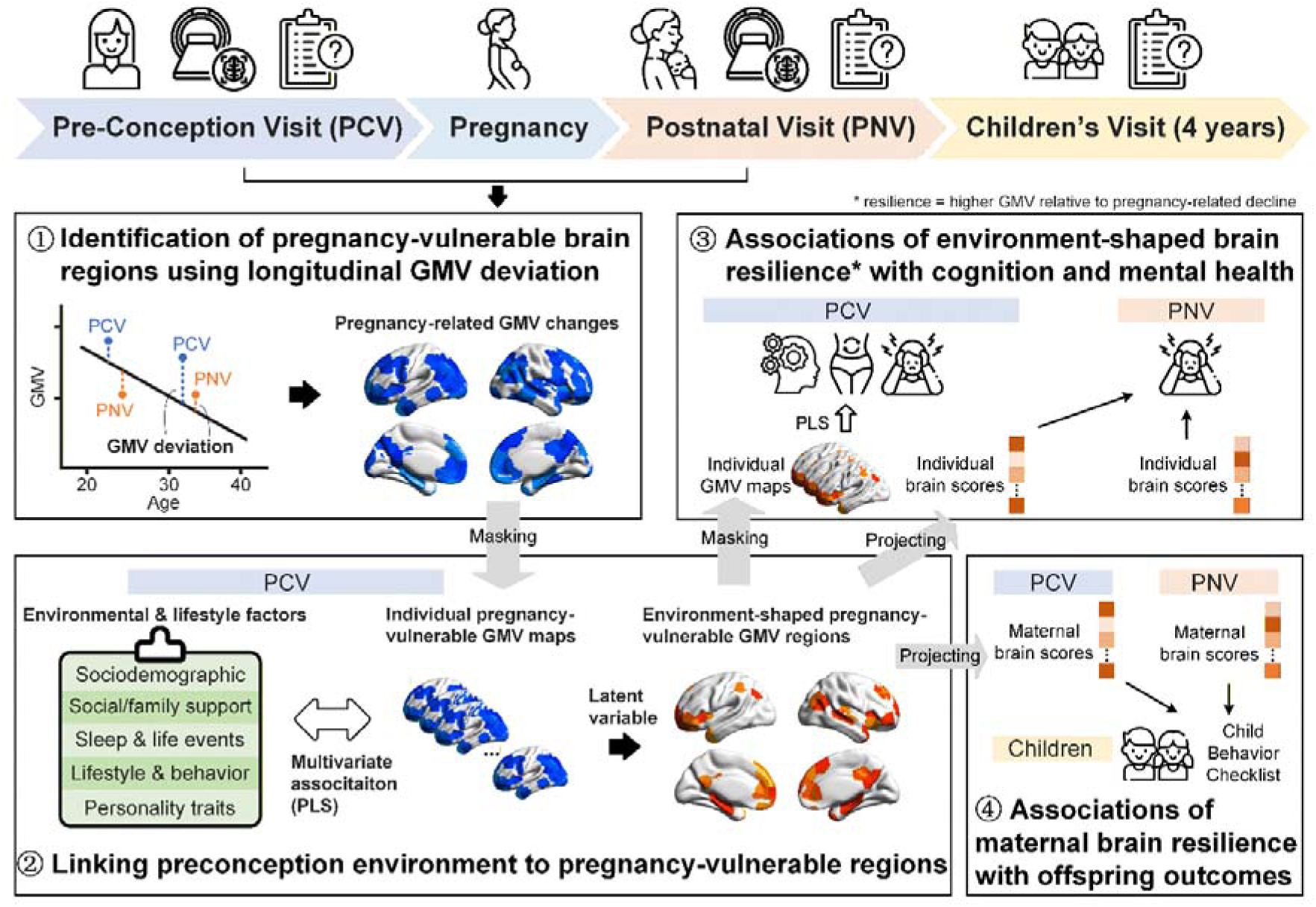
Study design. The longitudinal study timeline includes a pre-conception visit (PCV), postnatal visit (PNV) at 3 months and 6 months postpartum, and children’s assessment at 4 years of age. The analytical workflow comprised four steps: ① Using cross-sectional PCV data (N = 194), region-wise linear regression models were fitted to characterize normative age-related variation in gray matter volume (GMV). These age-based models were then applied to the 35 women with longitudinal MRI to compute GMV deviation (observed minus age-predicted GMV) at PCV and PNV. Paired tests of GMV deviation (PCV vs. PNV) identified pregnancy-vulnerable regions exhibiting significant pregnancy-related structural change. ② At PCV, partial least squares (PLS) analysis related 52 lifestyle, behavioral, sociodemographic, personality, and social/family support factors to GMV within pregnancy-vulnerable regions to environmentally associated structural variation and the contributing environmental factors. ③ To examine functional relevance, at PCV, the individual environment-shaped pregnancy-vulnerable GMV maps were associated with cognition, mental health, and metabolic outcomes using PLS analysis. Subsequently, individual GMV maps were projected onto the thresholded brain salience map derived from the GMV–environment PLS (see ②) to generate brain scores indexing structural resilience (i.e., relative preservation of GMV in pregnancy-vulnerable regions). These brain scores at PCV and PNV were then associated with mental health outcomes at PNV. ④ Finally, maternal brain scores at PCV and PNV were examined in relation to offspring socioemotional and behavioral outcomes (assessed by the Child Behavior Checklist) at 4 years to test cross-generational relevance.

### Pregnancy is associated with region-specific reductions in brain gray matter

We identified pregnancy-vulnerable brain regions by quantifying structural changes from preconception to postpartum while accounting for normative age-related variation. Age related regional gray matter volume (GMV) trajectories were modelled using all PCV participants, enabling the calculation of region-wise GMV deviation scores (observed minus age-predicted GMV) for longitudinal participants at both PCV and PNV (**Fig. 1** ①). Across 400 cortical and 14 subcortical regions of interest (ROIs), 93 regions showed significant pregnancy-related changes in GMV deviation from PCV to PNV following Holm–Bonferroni correction (corrected p < 0.05; **Fig. 2**). All significant regions exhibited lower GMV deviation at postpartum relative to preconception, indicating structural reductions beyond normative age-related expectations.

**Figure 2.**
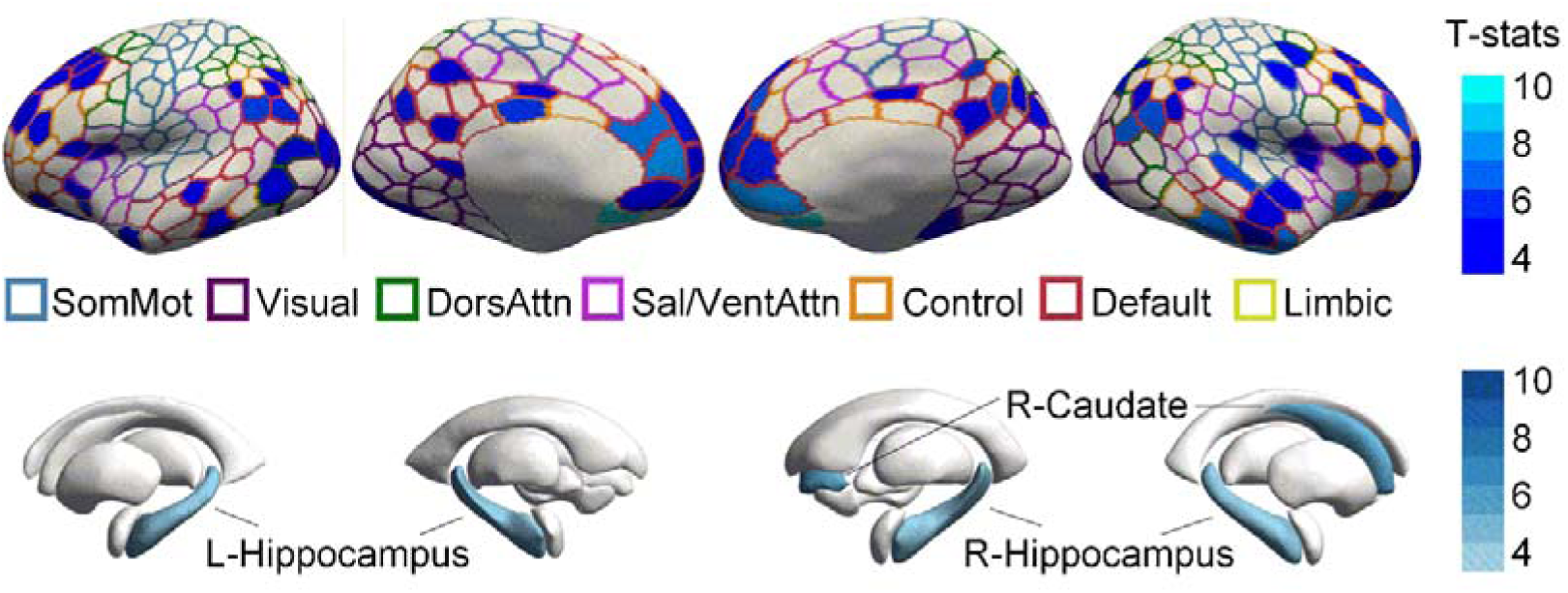
Brain regions showing significant pregnancy-related gray matter volume (GMV) reductions. Surface maps display the 93 cortical and subcortical regions in which GMV deviation (observed minus age-predicted GMV) significantly decreased from pre-conception (PCV) to 3-month postpartum (PNV), after Holm–Bonferroni correction (corrected p < 0.05) across 414 ROIs. Regions are shown on inflated cortical surfaces (lateral and medial views) and mapped to their functional network identities (SomMot = Somatomotor, Visual, DorsAttn = Dorsal attention, Sal/VentAttn = Salience/Ventral attention, Control, Default, Limbic). Subcortical effects were observed bilaterally in the hippocampus and in the right caudate. Color indicates t-statistics for paired comparisons (blue: PCV > PNV). No regions showed significant increases (PCV < PNV).

The pregnancy-related brain areas were widely distributed across large-scale cortical networks but showed clear functional specificity (**Fig. 2**). The default mode network exhibited the broadest involvement, with significant reductions across medial prefrontal, posterior cingulate/precuneus, inferior parietal, lateral temporal, and retrosplenial subdivisions in both hemispheres. Within the frontoparietal control network, vulnerable regions were distributed across lateral prefrontal, intraparietal, inferior parietal, and temporal control subdivisions. Limbic network involvement was also present, particularly in the orbitofrontal cortex and right temporal pole. In the dorsal attention network, reductions were observed in parieto–occipital, temporo–occipital, and superior parietal parcels. Several regions in the salience/ventral attention network, including fronto-insular, orbitofrontal, and frontal opercular areas, also showed GMV deviation reductions. Within the visual network, multiple extrastriate parcels showed significant reductions. Subcortically, the bilateral hippocampus and the right caudate nucleus showed significant GMV deviation reductions, consistent with the well-recognized plasticity of limbic–striatal circuits [35].

### Preconception lifestyle and psychosocial environments associated with GMV in pregnancy-vulnerable regions

We next examined whether preconception lifestyle and environmental factors contribute to the individual differences in preconception GMV within these pregnancy-vulnerable regions using a multivariate association approach (**Fig. 1**②).

A partial least squares (PLS) analysis relating GMV from the 93 pregnancy-vulnerable ROIs (**Fig. 2**) to 52 sociodemographic, lifestyle, behavioural, personality, and social/family support measures assessed at preconception (**Table 1**) resulted in a single significant latent variable (LV1) (permutation p < 0.001), explaining 38.7% of the covariance between pregnancy-vulnerable GMV and environmental factors. Thirty-seven ROIs showed stable contributions to this latent variable (|bootstrap ratio [BSR]| ≥ 2; **Fig. 3A**), indicating robust regional associations with the environmental pattern. These regions were primarily located within the default mode and frontoparietal control networks (medial/dorsal prefrontal cortex, inferior parietal lobe, posterior cingulate cortex, and temporoparietal junction), as well as salience and limbic regions (bilateral insula/operculum, orbitofrontal cortex) and subcortical structures (bilateral hippocampus, right caudate) (**Fig. 3 A**).

**Figure 3.**
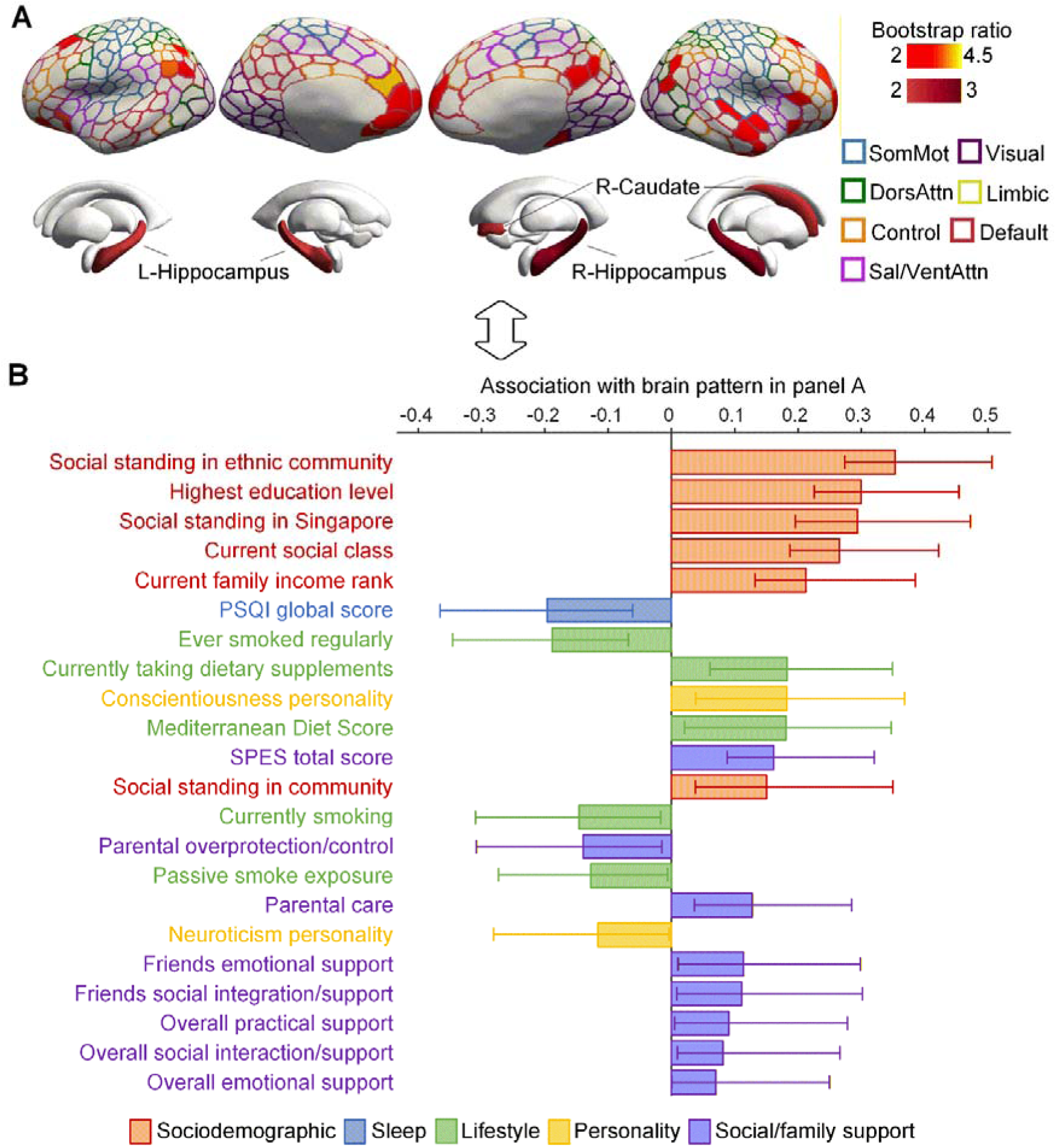
Lifestyle and environmental factors associated with pregnancy-vulnerable GMV at preconception. **A.** Cortical and subcortical regions contributing reliably (|BSR| ≥ 2) to the significant latent variable identified by PLS analysis linking lifestyle and environmental variables (Table 1) to GMV within pregnancy-vulnerable ROIs (Fig. 2). **B.** Environmental factors showing significant bootstrap-estimated contributions (95% confidence intervals not crossing zero). Positive bars indicate factors associated with higher GMV in pregnancy-vulnerable regions (e.g., higher social standing, greater educational attainment, higher income, higher Mediterranean Diet Score, greater conscientiousness, and stronger social support). Negative bars indicate factors associated with lower GMV, including poorer sleep quality, smoking history, current smoking, passive smoke exposure, and higher neuroticism.

**Table 1.**
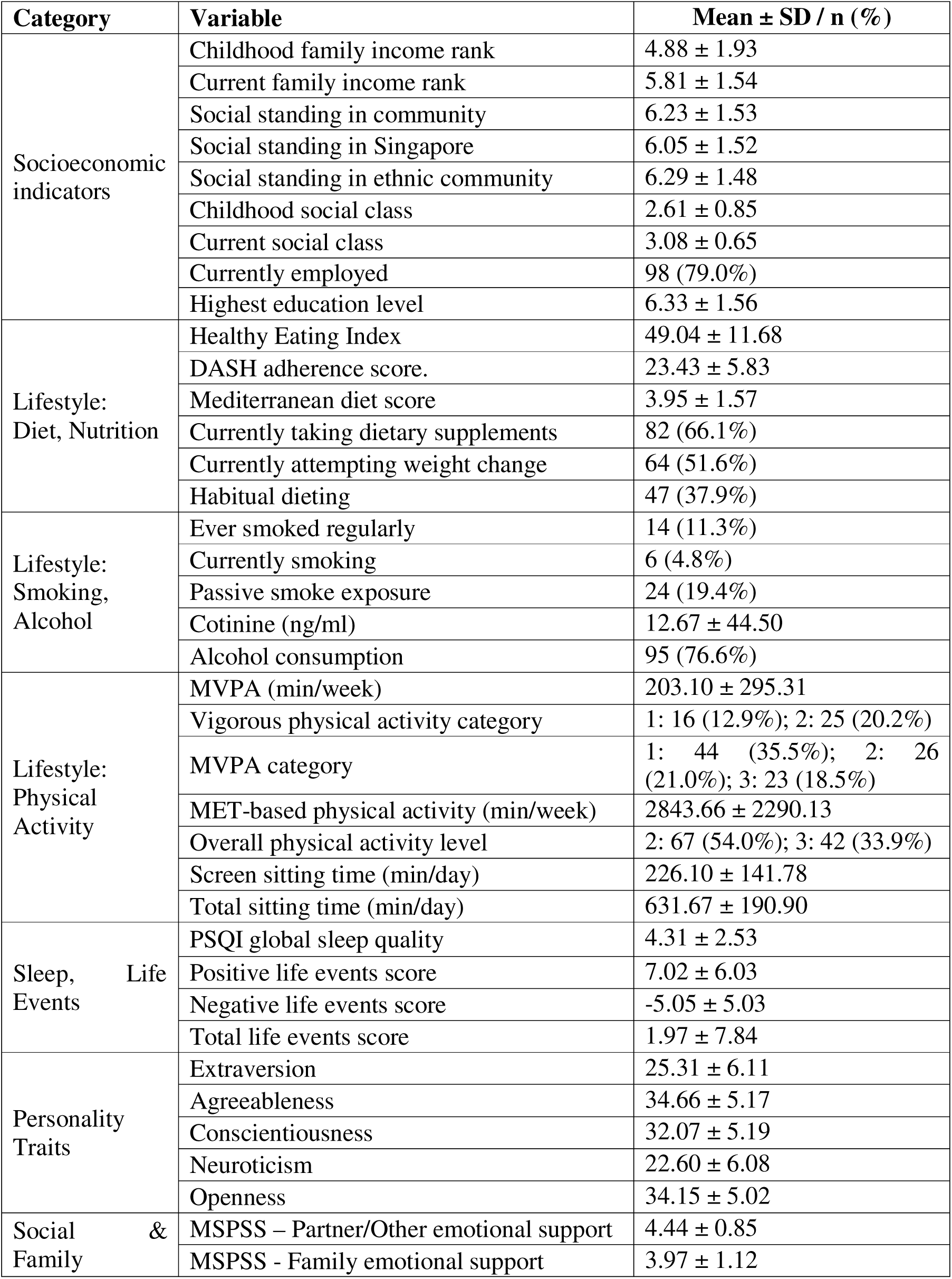
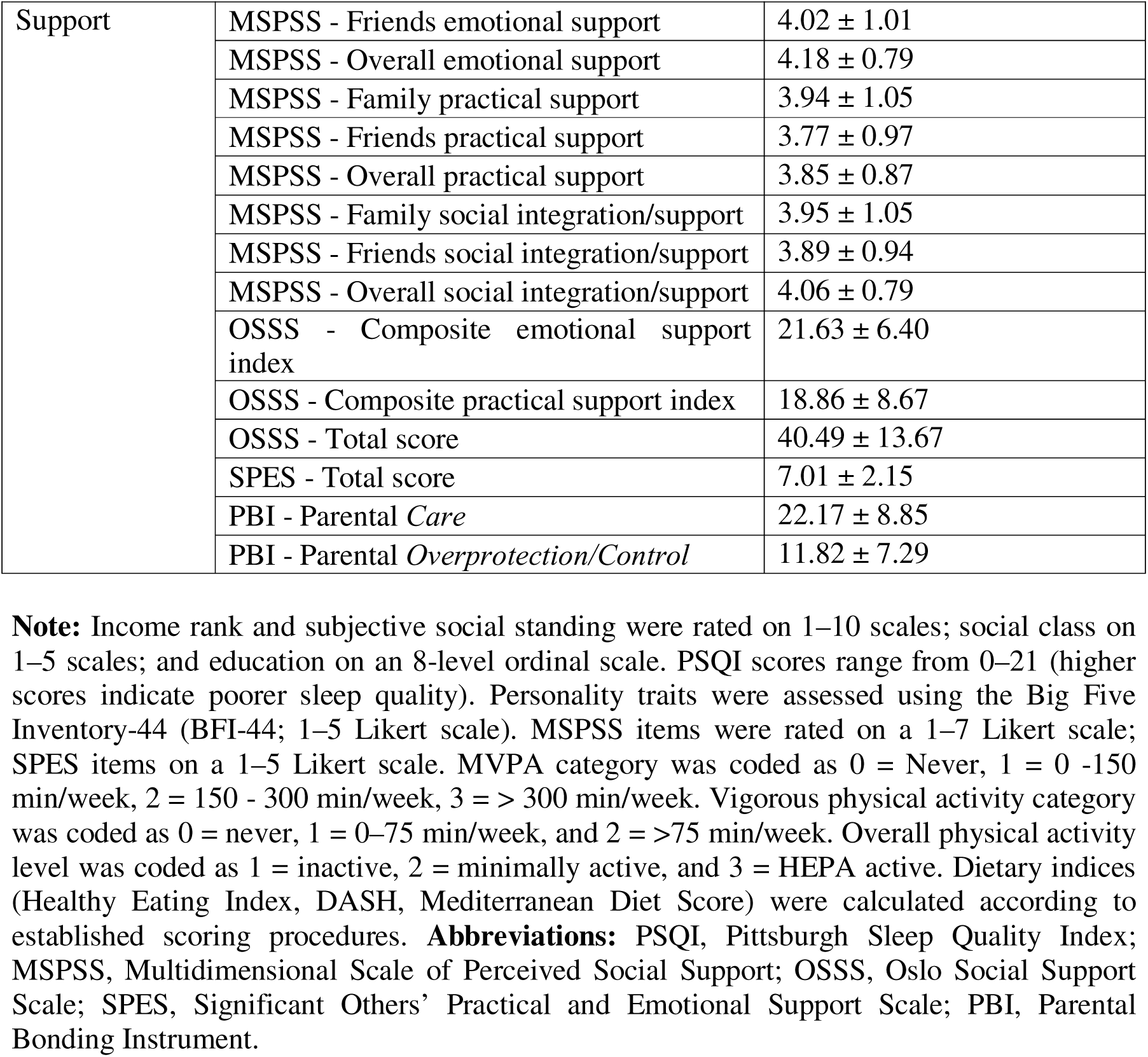
Preconception lifestyle and environmental measures in the PCV environment–GMV PLS sample (N = 124).

On the environmental side, 22 factors showed significant contributions, defined by bootstrap-derived 95% confidence intervals not crossing zero (**Fig. 3B**). Sociodemographic advantages showed the strongest positive associations with GMV, including greater educational attainment, higher social standing in Singapore and community, current social class, and higher current family income rank, indicating women with more favorable socioeconomic contexts exhibited greater structural robustness in brain regions later vulnerable to pregnancy-related reductions. Several lifestyle factors were also positively related to GMV, including higher Mediterranean diet score and active use of dietary supplements. In contrast, poorer sleep quality (higher PSQI score), regular smoking, current smoking, and passive smoke exposure were negatively associated with GMV, suggesting detrimental lifestyle exposures may weaken preconception structural resilience. Personality traits contributed meaningfully: higher conscientiousness was positively associated with GMV, whereas higher neuroticism showed negative associations, consistent with contrasting tendencies toward self-regulation vs. stress sensitivity. Multiple social and family support indices were also positively related to GMV, including emotional and instrumental support from partners, significant others and family/friends, overall social integration/support, and parental bonding (higher parental care, lower overprotection/control).

### Environment-shaped brain GMV vulnerability was associated with maternal cognition, mental health, and metabolism at preconception

We then tested whether GMV patterns shaped by these preconception environmental influences carries functional significance (**Fig. 1**③). At PCV, a PLS analysis relating GMV from the 37 environment-shaped pregnancy-vulnerable regions to 34 cognitive, mental health, and metabolic outcomes revealed a significant latent variable (permutation p < 0.001; LV1 explaining 46.9% of the covariance), indicating a robust multivariate brain–behavior association (**Fig. 4A**). Eleven brain regions contributed most strongly to this relationship, primarily within the default mode network (medial and dorsomedial prefrontal cortex, temporal regions), the frontoparietal control network (inferior parietal sulcus, ventrolateral prefrontal cortex), the salience network (anterior insula), and limbic/orbitofrontal and temporal–parietal areas.

**Figure 4.**
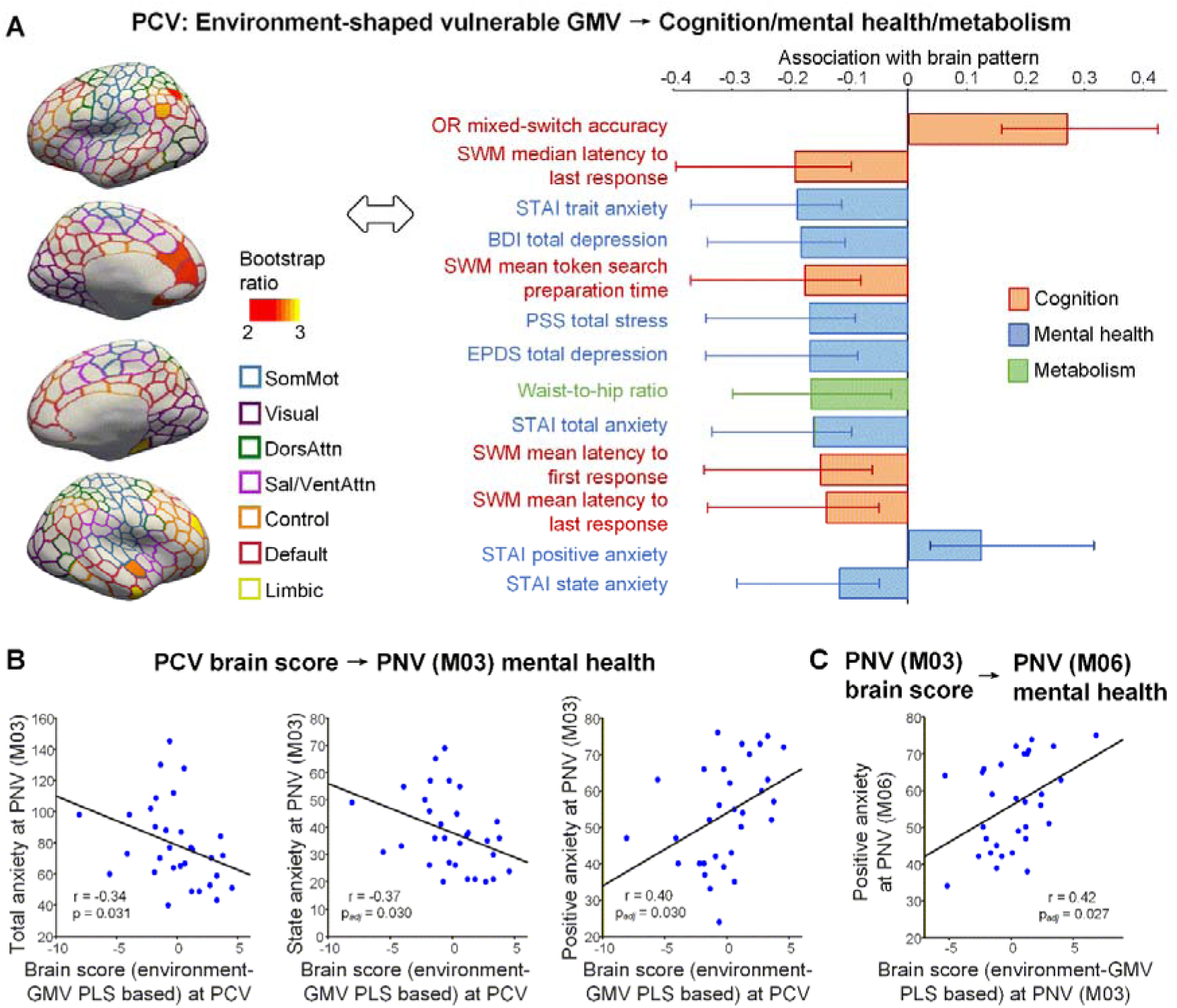
Associations between environment-shaped pregnancy-vulnerable GMV patterns and functional outcomes. **A.** Multivariate association (PLS) results relating GMV in environment-shaped pregnancy-vulnerable ROIs (|bootstrap ratios| ≥ 2) to cognitive, mental-health, and metabolic measures (95% CI not crossing zero) at PCV. **B.** Higher individualized brain scores (reflecting greater expression of the environment-shaped GMV pattern in Figure 3) at PCV predicted lower anxiety symptoms and higher positive-affect anxiety at PNV (M03). **C.** PNV M03 brain scores also predicted PNV M06 positive-affect anxiety symptoms. P_adj_ indicated FDR corrected p-values.

Higher GMV within this pattern was associated with better executive functioning (higher DCCS-OR mixed-switch accuracy indicating higher cognitive flexibility; shorter spatial working memory response latencies and token search preparation time), better mental health (lower STAI anxiety, BDI and EPDS depression scores, and perceived stress), and more favorable metabolic health (lower waist-to-hip ratio).

### Environment-shaped brain GMV vulnerability predicted postpartum mental health outcomes

We next tested whether this environmental-exposure-related brain resilience patterns predicted future postpartum health outcomes. We derived individualized brain scores at both PCV and PNV by projecting each participant’s GMV map onto the thresholded GMV–environment LV1 brain salience map (|BSR| ≥ 2; **Fig. 3A**). Higher brain scores reflected greater expression of the environment-linked structural pattern (i.e., relatively higher GMV within pregnancy-vulnerable regions). These scores were then examined in relation to postpartum mental health outcomes.

Higher PCV brain scores predicted more favorable emotional outcomes at 3 months postpartum (M03), including lower total anxiety (STAI total anxiety: r = –0.34, p = 0.031; STAI state anxiety: r = –0.37, p*_adj_*= 0.030; STAI positive affect/anxiety reduction: r = 0.40, p*_adj_* = 0.030; **Fig. 4B**) and showed a nominal association with lower anxiety at 6 months postpartum (STAI positive affect/anxiety reduction: r = 0.41, p = 0.040), but this effect did not survive FDR correction.

At the postpartum MRI visit (PNV), higher brain scores continued to index emotional resilience. The PNV M03 brain score was nominally associated with concurrent mental health at 3 months postpartum (STAI state anxiety: r = –0.26, p = 0.045; STAI positive affect/anxiety reduction: r = 0.30, p = 0.022). It also showed prospective associations with emotional outcomes at 6 months (M06) postpartum (STAI state anxiety: r = –0.30, p = 0.049), and the association with STAI positive affect/anxiety reduction remained significant after FDR correction (r = 0.42, p*_adj_* = 0.027; **Fig. 4C**).

### Environment-shaped GMV vulnerability predicted offspring mental health outcomes

Finally, we examined the intergenerational relevance of maternal brain structure by testing whether the identified environment-shaped pregnancy-vulnerable GMV patterns were associated with children’s socio-emotional and behavioral outcomes at age four (**Fig. 1**④).

At preconception (PCV), higher maternal brain scores reflecting greater expression of the environment-shaped pregnancy-related GMV resilience pattern (i.e., higher GMV values within the brain regions in **Fig. 3A**) were associated with better child mental health outcomes, including lower withdrawn behavior (r = -0.39, p = 0.006) and fewer depressive problems (r = -0.29, p = 0.034) (**Fig. 5A**).

**Figure 5.**
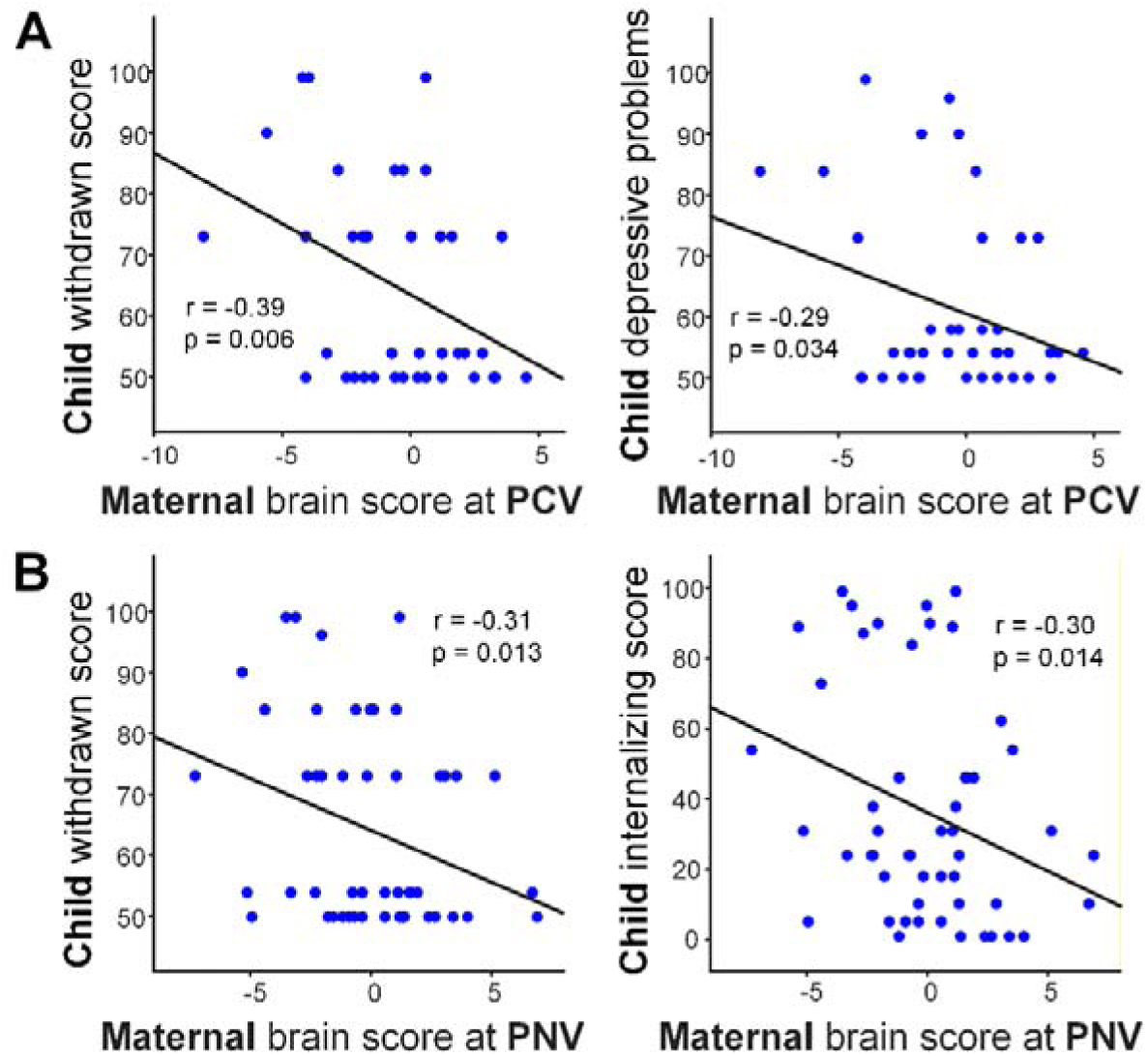
Associations between maternal environment-shaped brain resilience and children’s socio-emotional outcomes. **A.** Higher maternal brain scores at PCV (reflecting greater expression of the environment-shaped GMV pattern in Figure 3) were associated with lower CBCL withdrawn scores and depressive problem scores in children. **B.** Higher maternal brain scores at PNV were associated with lower withdrawn and internalizing problems in children.

At the postnatal visit (PNV, 3 months postpartum), this pattern remained predictive: higher maternal PNV brain scores were associated with reduced child withdrawn behavior (r = -0.31, p = 0.013) and fewer internalizing problems (r = -0.30, p = 0.014) (**Fig. 5B**).

## DISCUSSION

In this longitudinal preconception–postpartum–offspring neuroimaging study, we identified a distributed set of brain regions that undergo pregnancy-related structural remodeling, showed that modifiable preconception lifestyle and environmental factors are associated with structural resilience in these regions, and demonstrated that this environment-linked brain pattern prospectively predicts maternal cognitive, emotional, and metabolic outcomes, as well as children’s socio-emotional functioning. To our knowledge, this is the first study to (i) systematically evaluate how modifiable preconception environments relate to variability in pregnancy-related gray matter remodeling, and (ii) test whether this identified maternal structural patterns shaped by preconception environment index mental health across the pre- and postnatal periods and carry cross-generational relevance for child behavior. Together, these findings highlight the preconception period as a critical window for influencing maternal brain health and promoting positive outcomes for both mothers and their children.

### Pregnancy induces coordinated structural remodelling across distributed cortical–subcortical networks

Consistent with previous research demonstrating pregnancy-related reductions in GMV [5–9], we identified widespread structural alterations spanning cortical and subcortical systems. Vulnerable regions encompassed the default mode, frontoparietal control, salience, dorsal attention, and visual networks, as well as hippocampal and striatal structures. These networks support socio-emotional processing, executive control, stress regulation, and reward learning, all of which are essential for caregiving [11, 21, 22]. By correcting for age-related variation, our findings provide direct evidence that pregnancy is associated with coordinated remodeling beyond normative aging. This distributed pattern supports the notion that pregnancy triggers large-scale reorganization of cortico-limbic–striatal systems to facilitate maternal adaptation [1, 5, 36, 37].

### Lifestyle, socioeconomic, psychological, and social environments shape neural vulnerability before pregnancy

A central contribution of this study is the identification of modifiable preconception environmental factors associated with structural robustness in pregnancy-vulnerable regions. A single multivariate pattern linked higher preconception GMV to a constellation of favorable circumstances, including higher education, income, and perceived social standing; healthier dietary patterns; better sleep; avoidance of smoking and passive smoke exposure; stronger social support; and psychological resilience traits such as conscientiousness and lower neuroticism. These findings underscore that the maternal brain does not enter pregnancy as a blank slate. Instead, neural structure in regions later undergo pregnancy-related remodeling appears to be shaped by accumulated social, behavioral, and psychological exposures prior to conception, effectively determining the brain’s structural resilience.

This result aligns with broader evidence from developmental and lifespan neuroscience showing that socioeconomic conditions, health behaviors, sleep, and social environments influence cortical and limbic structure through pathways involving stress regulation, metabolic health, inflammation, and neuroplasticity [38–40]. Many of these factors are modifiable, identifying actionable targets for preconception health enhancement.

### Environment–shaped GMV vulnerability predicted maternal cognition, emotional health, and metabolic function

Pregnancy-related GMV vulnerability was not merely structural but functionally meaningful. At preconception, higher GMV in environment-shaped pregnancy-vulnerable regions was associated with better executive functioning, more favorable metabolic profiles, and lower levels of depression, anxiety, and perceived stress. These associations suggest that the neural systems undergoing pregnancy-related remodeling are already behaviorally relevant before conception. Importantly, this suggests that variability in these regions may represent a neural pathway through which modifiable lifestyle and environmental factors influence maternal cognitive, emotional, and metabolic functioning.

Crucially, preconception brain scores indexing greater structural robustness in regions undergoing pregnancy-related remodeling predicted emotional outcomes months after childbirth, indicating that structural patterns shaped by environmental factors may confer resilience to postpartum mental health difficulties. This finding offers a novel neural perspective on postpartum depression and anxiety, suggesting that vulnerability may be rooted not only in pregnancy-related changes but also in pre-existing neural organization shaped by lifestyle and environmental contexts [41].

### Environment–shaped maternal structural robustness predicts children’s socio-emotional development

We further demonstrated that maternal pregnancy-vulnerable GMV, particularly the component linked to maternal lifestyle and environmental factors, predicted children’s socio-emotional and behavioural outcomes at age four. These findings suggest that maternal neural health across pregnancy may shape the early caregiving environment, influence stress exposure, and affect mother–child interactions, thereby contributing to children’s socio-emotional development [22, 27, 42–44]. Given that these neural patterns were partly associated with modifiable lifestyle and environmental factors, preconception environments may indirectly relate to child socio-emotional outcomes through their association with maternal brain structure.

This cross-generational association aligns with emerging evidence that maternal brain structure and function contribute to maternal sensitivity, bonding, and emotional availability, which are key determinants of childhood socio-emotional trajectories [45–47]. Previous work in the same cohort showed that maternal stress at preconception influences neonatal brain microstructure and downstream offspring behaviour [48]. Our study extends this literature by showing that preconception neural characteristics, shaped by modifiable lifestyle and environmental factors, carry implications not only for maternal well-being but also for the next generation.

### Implications for preconception care and public health

Most existing research on maternal brain plasticity focuses on pregnancy or postpartum periods, overlooking the preconception stage—a window increasingly recognized as vital for lifelong maternal and child health [49, 50]. The preconception period captures baseline neural architecture before gestational biological stressors emerge, offering a unique opportunity to characterize the brain’s initial state and identify factors shaping susceptibility to later pregnancy-related remodeling.

Our findings highlight the preconception period as a critical window for promoting maternal neural resilience. Interventions that improve sleep, nutritional quality, social support, and socioeconomic stability, and reduce smoking and other harmful exposures, may strengthen structural robustness in brain networks that are particularly vulnerable to pregnancy-related structural change. Such improvements could in turn buffer against perinatal mental health difficulties and support healthier developmental environments for children.

Our results align with global recommendations that preconception health should be integrated into maternal and child health policy [51, 52]. The present findings extend this framework to the neural level, suggesting that supporting women before they conceive may have direct neurobiological and cross-generational benefits.

### Limitations and future directions

Several limitations should be noted. First, the postpartum imaging timepoint captures only early postpartum remodelling but does not reveal longer-term recovery trajectories. Future studies should include later postpartum assessments and more intensive longitudinal sampling across pregnancy and the postpartum period. Second, the longitudinal sample was modest. However, the inclusion of the preconception imaging substantially strengthens inference regarding pregnancy-related effects. Third, although we examined a wide range of environmental influences, additional factors such as hormonal profiles, stress biomarkers, sleep physiology, and paternal characteristics, may further contribute to variability in maternal brain structure and should be incorporated in future work. Fourth, children’s outcomes were parent-reported; integrating observational and neurobiological assessments will enhance precision. Fifth, the present findings are associative and do not establish causal relationships between environmental exposures, maternal brain structure, and offspring outcomes.

Future research should examine mechanistic pathways linking environmental exposures, maternal neural plasticity, perinatal mental health, and child development; incorporate richer physiological and hormonal measures; integrate multimodal MRI approaches (e.g., functional connectivity, white-matter microstructure); and test whether targeted preconception interventions can enhance neural resilience in pregnancy-vulnerable circuits.

## Conclusion

This study provides the first integrated evidence that the brain systems that undergo structural remodeling during pregnancy already show preconception vulnerability or resilience that is shaped by modifiable lifestyle and environmental factors. Preconception differences in grey matter volume pattern within these environment–shaped, pregnancy-vulnerable regions carry important implications for both maternal and child well-being. Together, these findings highlight the preconception period as a critical window for optimizing maternal brain health and supporting positive intergenerational outcomes.

## METHODS

### Participants

The present study used data from the Singapore S-PRESTO, a prospective preconception cohort designed to investigate early-life determinants of maternal and offspring health. Between February 2015 and October 2017, 1,032 Singaporean women of Chinese, Malay, or Indian ethnicity (or any combination thereof), aged 18-45 years and planning to conceive within the next 12 months, were recruited from KK Women’s and Children’s Hospital and the general population. Eligible participants intended to reside in Singapore for at least 5 years and were willing to provide written informed consent. Women were excluded if they were pregnant or breastfeeding at recruitment, had type 1 or type 2 diabetes; were taking systemic steroids, anticonvulsants, or antiviral medications for HIV/Hepatitis B/C within the prior month; had been attempting conception for more than 18 months; or had used oral or implanted contraception, intrauterine devices, or fertility medications within the prior month. Among recruited participants, 475 conceived singleton pregnancies within one year of enrollment, and 373 delivered liveborn infants who have been followed longitudinally into early childhood. Detailed cohort methods have been published previously [29]. The study received ethics approval from the SingHealth Centralised Institutional Review Board (reference 2014/692/D), and written informed consent was obtained from all participants.

The present analyses included women with structural MRI at preconception (PCV; n = 194) and/or 3 months postpartum (PNV; n = 61), including 35 longitudinal pairs. Child behavioral data at age four were available for 40 offspring of PCV mothers and 52 offspring of PNV mothers (**Fig. 1**).

### Lifestyle, environmental factors, metabolic indicators, and neuropsychological assessments

Lifestyle and environmental factors were assessed at the PCV (**Table 1**). Participants completed a comprehensive set of instruments capturing sociodemographic characteristics (income, education, subjective social status), lifestyle and behavioral factors including diet quality (Healthy Eating Index, DASH adherence, Mediterranean Diet Score, dietary supplement use), physical activity (moderate-to-vigorous activity time, walking time, Metabolic Equivalent of Task [MET]-based activity level, sedentary behavior), smoking and passive smoke exposure (self-reported smoking history, current smoking status, and salivary cotinine concentration), alcohol consumption, sleep quality (Pittsburgh Sleep Quality Index), life events (Life Experiences Survey), personality traits (Big Five Inventory-44), and social and family support (Multidimensional Scale of Perceived Social Support, Oslo Social Support Scale, Significant Others’ Practical and Emotional Support Scale, and Parental Bonding Instrument).

Cognitive, mental health, and metabolic measures were collected at the PCV (**Table 2**). Executive function was assessed using the Dimensional Change Card Sort task under emotion and orientation stimulus domains, respectively, yielding mean reaction times in pre-switch, post-switch, and mixed-switch conditions, as well as total accuracy in mixed-switch trials. Working memory was evaluated using the Cambridge Neuropsychological Test Automated Battery (CANTAB) Spatial Working Memory task, from which between-search errors, within-search errors, total errors, strategy scores, and several latency indices (mean latency to first response, mean and median latency to last response, and mean token search preparation time) were derived.

**Table 2.** Preconception cognition, mental health and metabolism measures in the PCV brain-outcome PLS sample (N = 128).

| Category | Variables / Measures | Mean ± SD |
| --- | --- | --- |
| **Cognitive Tasks: Executive Function** | DCCS-EM pre-switch mean reaction time (s) | 485.69 ± 117.11 |
|  | DCCS-EM post-switch mean reaction time (s) | 708.86 ± 243.05 |
|  | DCCS-EM mixed-switch mean reaction time (s) | 659.18 ± 193.97 |
|  | DCCS-EM mixed-switch accurate answers | 28.97 ± 1.28 |
|  | DCCS-OR pre-switch mean reaction time (s) | 285.29 ± 82.99 |
|  | DCCS-OR post-switch mean reaction time (s) | 699.21 ± 216.77 |
|  | DCCS-OR mixed-switch mean reaction time (s) | 677.24 ± 204.27 |
|  | DCCS-OR mixed-switch accurate answers | 29.19 ± 1.03 |
| **Cognitive Tasks: Working Memory** | CANTAB-SWM between-search errors | 19.61 ± 15.92 |
|  | CANTAB-SWM strategy score | 31.93 ± 5.54 |
|  | CANTAB-SWM mean latency to first response (s) | 1260.93 ± 498.71 |
|  | CANTAB-SWM mean latency to last response (s) | 21038.92 ± 3887.78 |
|  | CANTAB-SWM median latency to last response (s) | 19097.20 ± 3637.41 |
|  | CANTAB-SWM mean token search preparation time (s) | 817.45 ± 252.52 |
|  | CANTAB-SWM total errors | 20.10 ± 16.01 |
|  | CANTAB-SWM within-search errors | 1.13 ± 1.78 |
| **Mental health** | BDI total depression score | 8.46 ± 8.12 |
|  | EPDS total depression score | 7.91 ± 4.80 |
|  | STAI state anxiety score | 33.73 ± 10.50 |
|  | STAI trait anxiety score | 38.80 ± 10.73 |
|  | STAI total anxiety score | 72.54 ± 20.24 |
|  | STAI positive affect/anxiety-reducing items | 58.40 ± 11.25 |
|  | PSS total stress score | 16.13 ± 6.47 |
|  | Self-rated extent stress affected on health | 1.42 ± 0.89 |
|  | Self-rated stress in daily life | 1.58 ± 0.94 |
|  | GHQ total psychological distress score | 10.62 ± 5.89 |
|  | Need to Belong Scale total score | 33.05 ± 6.16 |
|  | General self-efficacy score | 15.45 ± 2.22 |
| **Metabolism** | Body Mass Index (kg/m<sup>2</sup>). | 23.64 ± 5.68 |
|  | Waist-to-Hip Ratio | 0.85 ± 0.07 |
|  | HbA1c level (%) | 5.12 ± 0.29 |
|  | HOMA-IR | 1.77 ± 1.67 |
|  | Fasting glucose from OGTT (mmol/L) | 4.85 ± 0.40 |
|  | 2-hour post-OGTT glucose (mmol/L) | 5.96 ± 1.66 |

**Note:** DCCS, Dimensional Change Card Sort; EM, Emotion task; OR, Orientation task; CANTAB, Cambridge Neuropsychological Test Automated Battery; SWM: Spatial Working Memory; BDI, Beck Depression Inventory; EPDS, Edinburgh Postnatal Depression Scale; STAI, State–Trait Anxiety Inventory; PSS, Perceived Stress Scale; GHQ, General Health Questionnaire; OGTT, Oral Glucose Tolerance Test. Reaction times and latencies are reported in seconds. Higher scores on BDI, EPDS, STAI (total, state, and trait anxiety), PSS, and GHQ indicate greater symptom severity. Higher scores on STAI positive affect/anxiety-reducing items indicate lower anxiety. HOMA-IR, Homeostatic Model Assessment of Insulin Resistance.

Mental health was assessed using a comprehensive standardized battery. Depressive symptoms were measured using the Beck Depression Inventory (BDI) and the Edinburgh Postnatal Depression Scale (EPDS). State anxiety, trait anxiety, total anxiety, and reverse-coded positive affect/anxiety-reduction scores were obtained from the State-Trait Anxiety Inventory (STAI). Perceived stress was captured using the Perceived Stress Scale (PSS), supplemented with two S-PRESTO–specific items evaluating (i) the extent to which stress has affected overall health and (ii) the amount of stress experienced in the previous four weeks. General psychological distress was quantified using the General Health Questionnaire–12 (GHQ-12). Additional psychological attributes included the Need to Belong Scale and the Generalized Self-Efficacy Scale, indexing desire for social acceptance and perceived coping capability.

Metabolic indicators included body mass index (BMI), waist-to-hip ratio, glycated hemoglobin (HbA1c), homeostatic model assessment of insulin resistance (HOMA-IR), fasting plasma glucose from oral glucose tolerance testing (OGTT), and 2-hour post-OGTT glucose concentrations.

Depression and anxiety were reassessed at the PNV at 3 months postpartum (M03) and 6 months postpartum (M06), using the same instruments administered at PCV, including the STAI, BDI, and EPDS (**Table 3**).

**Table 3.**
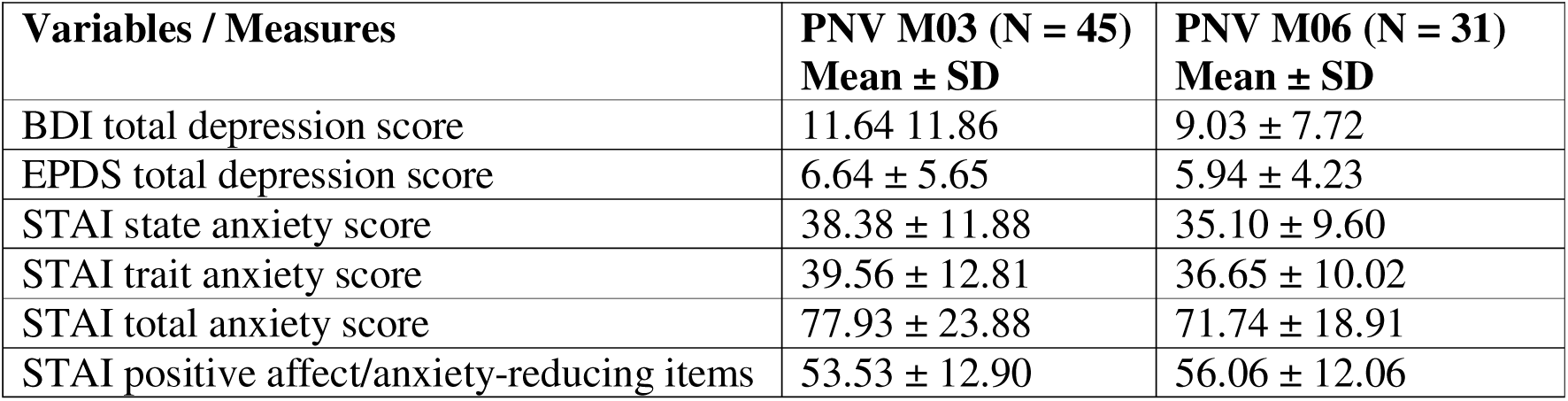
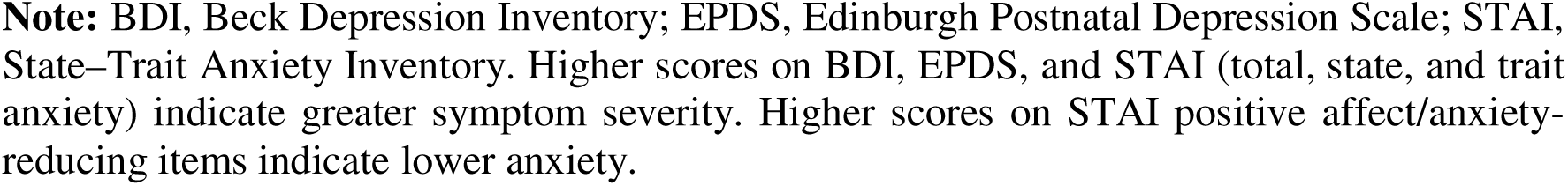
Postpartum mental health measures in the PNV MRI subsample.

| **Variables / Measures** | **PNV M03 (N = 45)**<br>**Mean ± SD** | **PNV M06 (N = 31)**<br>**Mean ± SD** |
| --- | --- | --- |
| BDI total depression score | 11.64 ± 11.86 | 9.03 ± 7.72 |
| EPDS total depression score | 6.64 ± 5.65 | 5.94 ± 4.23 |
| STAI state anxiety score | 38.38 ± 11.88 | 35.10 ± 9.60 |
| STAI trait anxiety score | 39.56 ± 12.81 | 36.65 ± 10.02 |
| STAI total anxiety score | 77.93 ± 23.88 | 71.74 ± 18.91 |
| STAI positive affect/anxiety-reducing items | 53.53 ± 12.90 | 56.06 ± 12.06 |

**Note:** BDI, Beck Depression Inventory; EPDS, Edinburgh Postnatal Depression Scale; STAI, State-Trait Anxiety Inventory. Higher scores on BDI, EPDS, and STAI (total, state, and trait anxiety) indicate greater symptom severity. Higher scores on STAI positive affect/anxiety-reducing items indicate lower anxiety.

Child socio-emotional and behavioral outcomes at age 4 years were evaluated using the Child Behavior Checklist for ages 1.5–5 (CBCL/1.5–5; **Table 4**). The CBCL provides standardized parent-reported ratings across multiple domains of child functioning and yields (i) syndrome scales, including Emotionally Reactive, Anxious/Depressed, Somatic Complaints, Withdrawn, Sleep Problems, Attention Problems, and Aggressive Behavior; (ii) DSM-oriented scales, including Depressive Problems, Anxiety Problems, Autism Spectrum Problems, Attention Deficit/Hyperactivity Problems, and Oppositional Defiant Problems; and (iii) broad-band composite scores for internalizing and externalizing behavior. Higher scores reflect greater behavioral or emotional difficulties.

**Table 4.** Child Behavior Checklist (CBCL/1½–5) measures in children of mothers from the PCV and PNV MRI samples.

| **Variables** | **Children of PCV mothers (N = 40)** | **Children of PNV mothers (N = 52)** |
| --- | --- | --- |
| Syndrome - Emotionally Reactive | 52.73 ± 5.11 | 52.21 ± 4.56 |
| Syndrome - Anxious/Depressed | 52.95 ± 5.27 | 52.65 ± 4.87 |
| Syndrome - Somatic Complaints | 53.78 ± 6.35 | 53.17 ± 6.10 |
| Syndrome - Withdrawn | 55.43 ± 8.60 | 55.06 ± 8.01 |
| Syndrome - Sleep Problems | 53.63 ± 4.59 | 53.87 ± 4.47 |
| Syndrome - Attention Problems | 52.43 ± 4.39 | 52.71 ± 4.60 |
| Syndrome - Aggressive Behavior | 52.20 ± 4.96 | 51.71 ± 4.35 |
| Internalizing Problem Score | 46.83 ± 11.65 | 45.40 ± 11.59 |
| Externalizing Problem Score | 44.00 ± 10.17 | 42.88 ± 10.37 |
| Total Problem Score | 45.23 ± 11.16 | 44.48 ± 11.12 |
| DSM - Depressive Problems | 53.53 ± 5.22 | 53.52 ± 4.92 |
| DSM - Anxiety Problems | 54.15 ± 4.76 | 53.73 ± 4.64 |
| DSM - Autism Spectrum Problems | 54.08 ± 8.51 | 53.29 ± 7.71 |
| DSM - Attention Deficit / Hyperactivity Problems | 51.63 ± 3.00 | 51.94 ± 3.47 |
| DSM - Oppositional Defiant Problems | 52.23 ± 4.25 | 51.92 ± 3.99 |

**Note:** Scores are presented as T-scores. Higher scores indicate higher risk.

### Image acquisition and preprocessing

High-resolution structural MRI data were acquired at PCV and PNV (M03) on a Siemens Prisma-fit 3T scanner using a standard head coil. A T1-weighted 3D MPRAGE sequence was collected with the following parameters: TR = 2000 ms, TE = 2.08 ms, TI = 877 ms, flip angle = 9°, voxel size = 1.0 × 1.0 × 1.0 mm³, 192 slices, and field of view = 192 mm. Acceleration was achieved using GRAPPA (acceleration factor = 2). GRAPPA acceleration (factor = 2) and Prescan Normalize were applied to optimize image quality.

We performed manual visual quality control on the raw T1-weighted images, rating each scan as good, pass, questionable, or fail based on the presence of motion artifacts. Only scans rated as good or pass were included for subsequent processing. Structural MRI data from both PCV and PNV were processed using FreeSurfer 6.0 following the standard “recon-all” pipeline. Briefly, non-brain tissue was removed using a hybrid watershed/surface deformation approach, followed by transformation to Talairach space, intensity normalization, and automated segmentation of subcortical white matter and deep gray matter structures. Cortical surfaces were generated via tessellation and topology correction, with white–gray matter and pial surfaces refined through intensity- and gradient-based surface deformation. Each participant’s cortical surface was then registered to a spherical atlas that aligns folding patterns across individuals.

For the 35 women with both PCV and PNV scans, we additionally applied FreeSurfer’s longitudinal recon-all pipeline, which creates an unbiased within-subject template to improve the reliability and sensitivity of longitudinal cortical measurements.

### Grey matter volume deviation estimation

Cortical gray matter volumes (GMV) were extracted using the Schaefer 400-parcel functional atlas, which was derived from resting-state fMRI from 1,489 young adults and optimized for correspondence between structural and functional organization [30]. In addition, gray matter volumes from 14 subcortical regions were obtained based on the Desikan–Killiany atlas [31]. This yielded 414 region-of-interest (ROI) GMV measures per participant at each visit.

To quantify pregnancy-related alterations in GMV, we first computed ROI-wise GMV deviation values using a linear regression–based predictive modeling approach adapted following the framework of Shen et al. [32, 33] (**Fig. 1** ①). First, age-related GMV variation was modeled using data from all women scanned at preconception (PCV; n = 194). For each of the 414 ROIs, a separate linear regression model was fitted:

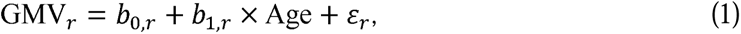

where *b*_O,r_ and *b*_l,r_ capture the age–GMV relationship at region r. These models established expected GMV values as a function of age, independent of pregnancy-related effects.

The resulting regression coefficients were then applied to the longitudinal subset of women with MRI scans at both PCV and 3 months postpartum (PNV; *n* = 35), to estimate age-expected GMV:

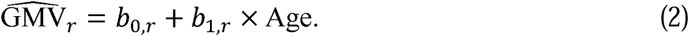

This provided individualized estimates of GMV expected solely from age-related variation.

Finally, for each ROI and visit in the longitudinal sample, GMV deviation was calculated as the difference between observed and age-expected GMV:

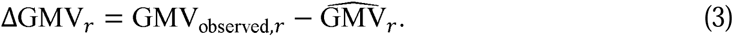

Negative deviations indicate lower-than-expected GMV (suggesting possible GMV reduction), whereas positive deviations reflect higher-than-expected GMV (suggesting relatively preserved or “younger-appearing” GMV).

### Statistical analysis

#### Identification of pregnancy-vulnerable GMV regions

For each ROI, paired t-tests were conducted to compare GMV deviation values between PCV and PNV (M03) within the 35 longitudinal participants. Multiple-comparison correction across all 414 ROIs was performed using the Holm–Bonferroni procedure (corrected p < 0.05). ROIs that remained significant after correction were classified as pregnancy-vulnerable GMV regions, reflecting brain areas showing systematic GMV alterations beyond age-expected variation during early postpartum.

#### Identification of lifestyle and environmental factors associated with pregnancy-vulnerable GMV at preconception

To examine whether preconception lifestyle and environmental factors were associated with inter-individual variability in GMV within regions showing pregnancy-related remodeling, we performed Partial Least Squares (PLS) analysis [34], a multivariate approach that identifies latent components maximizing the covariance between two sets of variables. Specifically, GMV values from the identified pregnancy-vulnerable regions were related to 52 lifestyle and environmental variables assessed at preconception, spanning sociodemographic characteristics, diet and nutrition, physical activity, smoking and alcohol use, sleep and life events, personality traits, and social/family support (**Table 1**; **Fig. 1** ②).

PLS analysis was conducted on the subset (N = 124) of women scanned at PCV who had complete imaging and behavioral data. The statistical significance of latent variables was assessed using 5,000 permutations, and the stability of feature contributions to the latent variables was evaluated using 5,000 bootstrap resamples. Reliable brain-region contributions were identified using a bootstrap ratio (BSR) threshold of |BSR| ≥ 2. The statistical reliability of environmental factors was defined by bootstrap-derived 95% confidence intervals for the regression weights that did not cross zero.

#### Prospective associations of environment-shaped pregnancy-vulnerable GMV patterns with cognitive, mental health, and metabolic outcomes

To evaluate whether environment-shaped brain resilience (i.e., greater structural preservation in pregnancy-vulnerable regions) may translate into benefits for cognition, mental health, and metabolic functioning, we examined associations between GMV in the environment-shaped pregnancy-vulnerable regions (i.e., ROIs identified from the previous GMV–environment PLS with |BSR| ≥ 2) and a set of outcomes measured at both preconception and postpartum (**Fig. 1** ③).

Specifically, at preconception, we conducted a PLS analysis between the GMV values of ROIs identified as environment-shaped pregnancy-vulnerable and the cognitive, mental health, and metabolic measures obtained at PCV (34 variables in **Table 2**). This analysis identified multivariate GMV–behavior relationships present before pregnancy.

Because the sample size was smaller at postpartum visits, a full multivariate PLS was not performed for other associations involving postpartum visits. Instead, for each participant, we derived an individualized brain score by projecting their raw GMV map onto the thresholded brain salience map (|BSR| ≥ 2) obtained from the GMV–environment PLS. Higher brain scores represent greater expression of higher GMV across environment-shaped pregnancy-vulnerable regions, indicating increased resilience to pregnancy-related structural change. We then tested whether this brain score predicted postpartum outcomes using Pearson’s correlations across the following intervals (**Fig. 1** ③): (i) PCV brain score → PNV M03 and M06 depression and anxiety symptoms; (ii) PNV M03 brain score → PNV M03 and M06 depression and anxiety symptoms. Multiple comparisons were controlled using false discovery rate (FDR) correction across symptom subdomains. These analyses tested whether structural vulnerability associated with environmental factors has downstream consequences for maternal emotional health across the perinatal transition.

### Prospective associations of environment-shaped pregnancy-vulnerable GMV pattern with offspring socio-emotional and behavioral outcomes

To evaluate intergenerational implications, we tested whether environment-shaped pregnancy-vulnerable GMV patterns were associated with children’s socio-emotional and behavioral outcomes at 4 years of age (**Fig. 1** ④). Pearson’s correlations were conducted between mothers’ brain scores (derived using the GMV–environment PLS brain salience map) at PCV and PNV and children’s CBCL/1.5–5 scores, including syndrome scales, DSM-oriented scales, and broad internalizing and externalizing composites. These analyses assessed whether environment-shaped maternal structural vulnerability was associated with offspring socio-emotional and behavioral development.

## Data availability statement

The raw data pertaining to this study has been kept in the database of the investigators’ group. Data access and sharing can be arranged by contacting the corresponding authors upon reasonable request. Data sharing will require a data transfer and collaborative agreement with the requestor.

## Acknowledgements

This study was supported by the Singapore National Medical Research Council (NMRC/OFLCG19May-0035, NMRC/CIRG/1485/2018, NMRC/CSA-SI/0007/2016, NMRC/MOH-00707-01, NMRC/CG/435 M009/2017-NUH/NUHS, CIRG21nov-0007, HLCA23Feb-0004 and OFYIRG23jul-0010), RIE2020 AME Programmatic Fund from A*STAR, Singapore (No. A20G8b0102), Ministry of Education (MOE-T2EP40120-0007 & T2EP2-0223-0025, MOE-T2EP20220-0001), and Yong Loo Lin School of Medicine Research Core Funding, National University of Singapore, Singapore.

## References

1. Servin-Barthet, C., et al., The transition to motherhood: linking hormones, brain and behaviour. Nature Reviews Neuroscience, 2023. 24(10): p. 605–619.

2. Ghahremani, T., et al., Women’s mental health services and pregnancy: a review. Obstetrical & gynecological survey, 2022. 77(2): p. 122–129.

3. Dye, C., K.M. Lenz, and B. Leuner, Immune system alterations and postpartum mental illness: evidence from basic and clinical research. Frontiers in global women’s health, 2022. 2: p. 758748.

4. Orchard, E.R., H.J. Rutherford, A.J. Holmes, and S.D. Jamadar, Matrescence: lifetime impact of motherhood on cognition and the brain. Trends in Cognitive Sciences, 2023. 27(3): p. 302–316.

5. Hoekzema, E., et al., Pregnancy leads to long-lasting changes in human brain structure. Nature neuroscience, 2017. 20(2): p. 287–296.

6. Servin-Barthet, C., et al., Pregnancy entails a U-shaped trajectory in human brain structure linked to hormones and maternal attachment. Nature Communications, 2025. 16(1): p. 730.

7. Pritschet, L., et al., Neuroanatomical changes observed over the course of a human pregnancy. Nature Neuroscience, 2024. 27(11): p. 2253–2260.

8. Hoekzema, E., et al., Mapping the effects of pregnancy on resting state brain activity, white matter microstructure, neural metabolite concentrations and grey matter architecture. Nature Communications, 2022. 13(1): p. 6931.

9. Niu, Y., et al., Longitudinal investigation of neurobiological changes across pregnancy. Communications Biology, 2025. 8(1): p. 82.

10. Kim, P., A.J. Dufford, and R.C. Tribble, Cortical thickness variation of the maternal brain in the first 6 months postpartum: associations with parental self-efficacy. Brain Structure and Function, 2018. 223(7): p. 3267–3277.

11. Barba-Müller, E., S. Craddock, S. Carmona, and E. Hoekzema, Brain plasticity in pregnancy and the postpartum period: links to maternal caregiving and mental health. Archives of women’s mental health, 2019. 22(2): p. 289–299.

12. Dufford, A.J., G. Patterson, and P. Kim, Longitudinal neuroanatomical increases from early to one-year postpartum. Brain Structure and Function, 2024. 229(9): p. 2479–2492.

13. Rehbein, E., et al., Pregnancy and brain architecture: associations with hormones, cognition and affect. Journal of neuroendocrinology, 2022. 34(2): p. e13066.

14. Zimmerman, M.E., et al., The relationship between frontal gray matter volume and cognition varies across the healthy adult lifespan. The American journal of geriatric psychiatry, 2006. 14(10): p. 823–833.

15. Schienle, A., F. Ebner, and A. Schäfer, Localized gray matter volume abnormalities in generalized anxiety disorder. European archives of psychiatry and clinical neuroscience, 2011. 261(4): p. 303–307.

16. Aleknaviciute, J., et al., Long-term association of pregnancy and maternal brain structure: the Rotterdam Study. European Journal of Epidemiology, 2022. 37(3): p. 271–281.

17. de Lange, A.M.G., et al., *The maternal brain: Region*l*specific patterns of brain aging are traceable decades after childbirth*. Human brain mapping, 2020. 41(16): p. 4718–4729.

18. Lucente, M. and J. Guidi, Allostatic load in children and adolescents: a systematic review. Psychotherapy and Psychosomatics, 2023. 92(5): p. 295–303.

19. Zhao, Y., et al., The brain structure, immunometabolic and genetic mechanisms underlying the association between lifestyle and depression. Nature Mental Health, 2023. 1(10): p. 736–750.

20. Tian, Y.E., J.H. Cole, E.T. Bullmore, and A. Zalesky, Brain, lifestyle and environmental pathways linking physical and mental health. Nature Mental Health, 2024. 2(10): p. 1250–1261.

21. Cárdenas, E.F., A. Kujawa, and K.L. Humphreys, Neurobiological changes during the peripartum period: implications for health and behavior. Social cognitive and affective neuroscience, 2020. 15(10): p. 1097–1110.

22. Graham, A.M., et al., Effects of maternal psychological stress during pregnancy on offspring brain development: considering the role of inflammation and potential for preventive intervention. Biological Psychiatry: Cognitive Neuroscience and Neuroimaging, 2022. 7(5): p. 461–470.

23. Barrett, J. and A.S. Fleming, Annual research review: All mothers are not created equal: Neural and psychobiological perspectives on mothering and the importance of individual differences. Journal of Child Psychology and Psychiatry, 2011. 52(4): p. 368–397.

24. Abraham, E., T. Hendler, O. Zagoory-Sharon, and R. Feldman, Network integrity of the parental brain in infancy supports the development of children’s social competencies. Social cognitive and affective neuroscience, 2016. 11(11): p. 1707–1718.

25. Disner, S.G., C.G. Beevers, E.A. Haigh, and A.T. Beck, Neural mechanisms of the cognitive model of depression. Nature Reviews Neuroscience, 2011. 12(8): p. 467–477.

26. Shin, L.M. and I. Liberzon, The neurocircuitry of fear, stress, and anxiety disorders. Neuropsychopharmacology, 2010. 35(1): p. 169–191.

27. Wu, Y., J. De Asis-Cruz, and C. Limperopoulos, Brain structural and functional outcomes in the offspring of women experiencing psychological distress during pregnancy. Molecular psychiatry, 2024. 29(7): p. 2223–2240.

28. Wu, Y., et al., Association of elevated maternal psychological distress, altered fetal brain, and offspring cognitive and social-emotional outcomes at 18 months. JAMA Network Open, 2022. 5(4): p. e229244–e229244.

29. Loo, E.X.L., et al., Cohort profile: Singapore preconception study of long-term maternal and child outcomes (S-PRESTO). European journal of epidemiology, 2020. 36(1): p. 129.

30. Schaefer, A., et al., Local-global parcellation of the human cerebral cortex from intrinsic functional connectivity MRI. Cerebral cortex, 2018. 28(9): p. 3095–3114.

31. Desikan, R.S., et al., An automated labeling system for subdividing the human cerebral cortex on MRI scans into gyral based regions of interest. Neuroimage, 2006. 31(3): p. 968–980.

32. Shen, X., et al., *Using connectome-based predictive modeling to predict individual behavior from brain connectivity*. nature protocols, 2017. 12(3): p. 506–518.

33. Sun, H., et al., Brain age prediction and deviations from normative trajectories in the neonatal connectome. Nature communications, 2024. 15(1): p. 10251.

34. Krishnan, A., L.J. Williams, A.R. McIntosh, and H. Abdi, Partial Least Squares (PLS) methods for neuroimaging: a tutorial and review. Neuroimage, 2011. 56(2): p. 455–475.

35. Kreitzer, A.C. and R.C. Malenka, Striatal plasticity and basal ganglia circuit function. Neuron, 2008. 60(4): p. 543–554.

36. Kim, P., L. Strathearn, and J.E. Swain, The maternal brain and its plasticity in humans. Hormones and behavior, 2016. 77: p. 113–123.

37. Swain, J.E., The human parental brain: in vivo neuroimaging. Progress in Neuro-Psychopharmacology and Biological Psychiatry, 2011. 35(5): p. 1242–1254.

38. McEwen, B.S. and P.J. Gianaros, Central role of the brain in stress and adaptation: links to socioeconomic status, health, and disease. Annals of the New York Academy of Sciences, 2010. 1186(1): p. 190–222.

39. Moussiopoulou, J. and P. Falkai, *The common biological and social bases for physical and mental disorders: focus on inflammation and neuroendocrine system*, in Comorbidity between mental and physical disorders: Identification, management and treatment. 2025, Springer. p. 61–87.

40. Davidson, R.J. and B.S. McEwen, Social influences on neuroplasticity: stress and interventions to promote well-being. Nature neuroscience, 2012. 15(5): p. 689–695.

41. Wang, Y., Y. Du, J. Li, and C. Qiu, Lifespan intellectual factors, genetic susceptibility, and cognitive phenotypes in aging: implications for interventions. Frontiers in Aging Neuroscience, 2019. 11: p. 129.

42. Shi, Y., et al., Longitudinal association between maternal psychological stress during pregnancy and infant neurodevelopment: the moderating effects of responsive caregiving. Frontiers in Pediatrics, 2022. 10: p. 1007507.

43. Tadanki, D., et al., Comprehensive Review of the Impact of Maternal Stress on Fetal Development. Pediatric Discovery, 2025: p. e70004.

44. Pan, B., et al., The impact of maternal parenting stress on early childhood development: the mediating role of maternal depression and the moderating effect of family resilience. BMC psychology, 2025. 13(1): p. 277.

45. Cainelli, E., L. Vedovelli, and P. Bisiacchi, The mother–child interface: A neurobiological metamorphosis. Neuroscience, 2024. 561: p. 92–106.

46. Nehls, S., et al., Time-sensitive changes in the maternal brain and their influence on mother-child attachment. Translational Psychiatry, 2024. 14(1): p. 84.

47. Gholampour, F., M.M. Riem, and M.I. van den Heuvel, Maternal brain in the process of maternal-infant bonding: Review of the literature. Social neuroscience, 2020. 15(4): p. 380–384.

48. Chan, S.Y., et al., Neonatal nucleus accumbens microstructure modulates individual susceptibility to preconception maternal stress in relation to externalizing behaviors. Journal of the American Academy of Child & Adolescent Psychiatry, 2024. 63(10): p. 1035–1046.

49. Stephenson, J., et al., Before the beginning: nutrition and lifestyle in the preconception period and its importance for future health. The Lancet, 2018. 391(10132): p. 1830–1841.

50. Fleming, T.P., et al., Origins of lifetime health around the time of conception: causes and consequences. The Lancet, 2018. 391(10132): p. 1842–1852.

51. Cassinelli, E.H., et al., Preconception health and care policies, strategies and guidelines in the UK and Ireland: a scoping review. BMC Public Health, 2024. 24(1): p. 1662.

52. Mason, E., et al., Preconception care: advancing from ‘important to do and can be done’to ‘is being done and is making a difference’. Reproductive health, 2014. 11(Suppl 3): p. S8.

